# Peripheral Mitochondrial Energetics are Associated with Cortical Neurophysiological Alterations in Alzheimer’s Disease

**DOI:** 10.64898/2026.03.25.26349329

**Authors:** Sean L. Kriwokon, Santiago I. Flores-Alonso, Brianne A. Kent, Tony W. Wilson, Rachel K. Spooner, Alex I. Wiesman

**Author notes:** Corresponding author, Alex I. Wiesman, PhD, Department of Biomedical Physiology & Kinesiology, Simon Fraser University, Burnaby, British Columbia.

## Abstract

Alzheimer’s disease is associated with both mitochondrial dysfunction and altered neurophysiological signalling. Peripheral measures of mitochondrial respiration have been established as effective predictors of mitochondrial function in the healthy brain, and more recently, of altered brain signalling in clinical groups. Here, we sought to assess whether peripheral mitochondrial energetics are associated with altered neural signalling in Alzheimer’s disease. We collected task-free magnetoencephalography (MEG) from individuals on the Alzheimer’s disease continuum (69.21 [6.91] years; n = 38) and cognitively normal older adults (72.20 [4.73] years; n = 20). Each participant also provided a blood sample for analysis of mitochondrial respiration using the Seahorse XF96 Analyzer. We used region-wise linear models to test the relationship between ATP-linked mitochondrial respiration and Alzheimer’s disease-associated neurophysiological changes. We found that mitochondrial respiration linked to ATP production is associated with altered alpha and theta band cortical rhythms in Alzheimer’s disease (α: *p*_FDR_ < 0.05, *r* = -0.7; θ: *p*_FDR_ < 0.05, *r* = -0.6). We then tested colocalization of mitochondria-neurophysiological relationships with a human brain atlas of respiratory capacity and found that brain regions with lower mitochondrial respiratory capacity exhibit a stronger relationship between aperiodic signalling and peripheral ATP-linked respiration (*p*_FDR_ = 0.003, *r* = 0.35). Our findings suggest that peripheral blood measures of mitochondrial function can offer insight into the neurophysiological alterations associated with energetic changes in Alzheimer’s disease and warrant further investigation into the translational potential of joint neuronal-mitochondrial markers of neurological diseases of aging.

## Introduction

Alzheimer’s disease (AD) is a neurodegenerative disorder characterized by the accumulation of amyloid-β plaques and hyper-phosphorylated tau tangles in the brain, accompanied by neuronal degradation and eventually leading to cognitive decline (1). The pathological build-up of these plaques and tangles is associated with both mitochondrial dysfunction (2) and altered cortical neurophysiological signalling (3). Evidence suggests that mitochondrial dysfunction impacts neurophysiological activity (4); however, the relationships between mitochondrial dysfunction and the cortical neurophysiological alterations observed in AD are unknown.

The presence of mitochondrial dysfunction in AD is well established, yet the direction of such effects appears to differ between studies of human patients and those using cell culture or rodent models (5). Non-human models of AD exhibit neuronal hypometabolism (6) and reduced mitochondrial respiration (7–9) compared to control levels. This mitochondrial dysfunction leads to increased accumulation of amyloid-β proteins (10), while interactions between Aβ, tau, and VDAC1 (mitochondrial voltage-gated ion channel) then contribute to worsened mitochondrial dysfunction (11). Importantly, the strongest genetic risk factor for sporadic AD (i.e., APOE4 allele; (12)) is associated with mitochondrial impairment in astrocytes (13), although human studies of mitochondrial function in clinical-stage AD often indicate that respiratory capacity is increased (14–17) and induced pluripotent stem cell (iPSC)-derived neurons from human patients express increased levels of the mitochondrial proteins involved in oxidative phosphorylation (18). Since non-human models of AD cannot fully recreate later-stage disease processes, this inconsistency may signal a non-linear trajectory of mitochondrial energetic changes along the AD continuum, mirroring alterations seen in neuronal activity (19,20).

Measuring mitochondrial function in the brains of living persons is invasive and not generally possible, making research into the neural effects of mitochondrial dysfunction in patients with AD difficult. Thus, minimally invasive methods of predicting mitochondrial function in the brain are needed to explore these effects. Peripheral mitochondria taken from whole blood samples have been identified as an accurate and relatively non-invasive method for predicting mitochondrial function in the brains of nonhuman primates (21), and can provide meaningful information relating to mitochondrial alterations in individuals with dementia (17).

Macro-scale neurophysiological signalling is also altered in AD, and particularly in response to the accumulation of Alzheimer’s proteinopathy. Task-free neuroimaging studies have shown that individuals with clinical-stage AD display aberrant patterns of neurophysiological ‘slowing’ characterized by a decrease in high frequency alpha (8-12 Hz) and beta (13-30 Hz) power accompanied by an increase in slower frequency theta (4-8 Hz) and delta (0.5-4 Hz) power (22). This neurophysiological slowing has been associated with proteinopathy and cognitive decline (3,20,23). Most literature pertaining to neurophysiological alterations in AD assesses periodic components of neural signaling, however brain signalling is not entirely periodic. More recently, emphasis is being placed on the importance of aperiodic components in normal brain function, with deviations indicating pathology. Altered aperiodic signalling has been associated with several neurological disorders (24–27), but findings regarding aperiodic neurophysiology in Alzheimer’s disease are inconsistent (28). Amongst positive findings, aperiodic parameters have helped researchers to differentiate between AD and frontotemporal dementia (29), and appear to track biophysical changes in excitation and inhibition in AD (30,31).

Converging evidence suggests that neurophysiological alterations in AD might be associated with mitochondrial dysfunction. Neuronal signalling is inherently ATP-hungry (32), and the emergence of periodic signalling is dependent on energy availability (33). Periodic signalling is also associated with mitochondrial dysfunction in other clinical disorders and *in vitro* models. For example, children with mitochondrial encephalomyopathies commonly exhibit slowing in alpha band activity (34). Spooner et al. (35) found evidence that mitochondrial dysfunction is correlated with pathological beta oscillations in people living with HIV. *In vitro* studies suggest that mitochondrial function is important for modulating neural oscillations via inhibitory inter-neuronal pools (36). These inhibitory interneurons produce high frequency activity and require a large, constant supply of ATP from mitochondria to maintain concentration gradients (37). Such interneurons are responsible for inhibitory drive on pyramidal cells which is crucial for gamma oscillations (37).

In sum, multiple lines of evidence support a possible link between mitochondrial function and multispectral neurophysiological signalling across disease states, although the mechanisms and processes remain poorly understood. To directly probe the relationship between mitochondrial energetics and altered cortical neurophysiology in AD, we collected peripheral blood samples and task-free magnetoencephalography (MEG) data from a group of biomarker-confirmed individuals on the AD continuum, as well as a group of cognitively normal older adults for comparison. The peripheral blood samples were used to estimate mitochondrial function with a Seahorse XF96 Analyzer, and the MEG data underwent source imaging and spectral parameterization to measure cortical neurophysiological signalling. We hypothesized that variability in peripheral mitochondrial function would be correlated with the region-specific alterations in cortical neurophysiological signalling that are characteristic of AD.

## Methods

### Participants

The Dynamic Mapping of Alzheimer’s disease Pathology (DMAP) study (38) recruited participants with aMCI and probable AD for collection of a dense battery of multimodal neuroimaging, neuropsychological testing, blood biomarker, and questionnaire data. Forty-four participants were referred from a memory disorders clinic and screened for inclusion in the AD continuum group. A fellowship trained neurologist determined that each of these participants had either aMCI or mild probable AD using standard clinical criteria (39), and whole-brain quantitative Aβ positron emission tomography (PET; see *Florbetapir ^18^F positron emission tomography* below). The final sample was n = 38 (aMCI: N = 18, probable AD: N = 20). The six excluded patients included one who withdrew due to safety concerns related to COVID-19, another who was excluded due to a major incidental finding, and four who were found to be Aβ negative on their PET scan.

Twenty additional older adults who reported no subjective cognitive concerns were screened for inclusion in this study. These adults were included for comparison and standardization of the AD continuum group relative to an analogous group of cognitively normal participants. A PET scan within the five years leading up to the study confirmed that 19 of these participants were Aβ negative. One participant did not undergo PET imaging but was included after performing exceptionally well on all neuropsychological tests. The 19 Aβ negative participants were previously enrolled in an unrelated clinical trial which confirmed that they were biomarker-negative. These participants underwent detailed neuropsychological assessments and did not exhibit any cognitive disturbances.

Exclusionary criteria for both groups included: medical illness affecting central nervous system (CNS) function, diagnosis of any neurological disorder other than AD, moderate or severe depression (Geriatric Depression Scale ≥ 10), current substance abuse, or history of head trauma. Demographic factors including highest level of education (*t* = 1.44, *p* = 0.156) and sex (χ^2^ = 0.84, *p* = 0.360) were matched across the two groups, but the groups differed in age (*t* = 2.02, *p* = 0.048) such that participants in the AD continuum group were younger than those in the cognitively normal (CN) group. Age was included as a nuisance covariate in all statistical analyses alongside highest level of education. Detailed clinical profiles and demographic group comparisons can be found in Table 1.

**Table 1.**
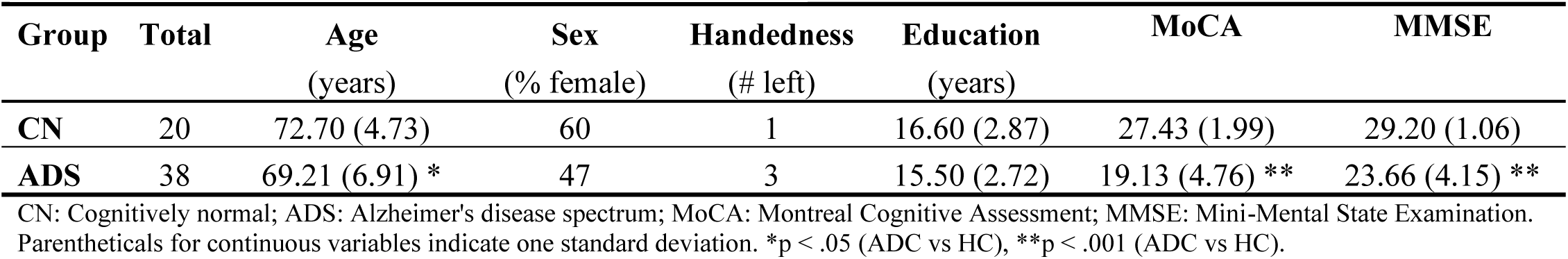
Participant demographics and cognitive profiles.

Each participant provided written informed consent after receiving a detailed description of the study. Participant informants were present for each participant on the AD continuum to ensure participant comfort throughout the course of the study as well as to complete certain questionnaires. Participant informants also provided written informed consent before taking part in the study. For participants with a questionable capacity to consent, informed assent was obtained from the research participant in addition to informed consent from a legally authorized representative to ensure that the interests of all participants were represented. The Institutional Review Board at the University of Nebraska Medical Center reviewed this investigation and granted approval. All research protocols complied with the Declaration of Helsinki.

### Neuropsychological Testing

After screening, and in collaboration with participant informants (spouse or child of participant) for individuals with AD or aMCI, the Functional Activities Questionnaire (FAQ) (40) was used to measure instrumental activities of daily living (iADL). The Quick Dementia Rating System (QDRS) (41) measured dementia severity and behavioural symptoms. The Montreal Cognitive Assessment (MoCA) (42) and Mini-mental State Examination (MMSE) (43) were used to measure general cognitive status. More details regarding the neuropsychological tests and questionnaires administered can be found in previous publications (23,38,44).

### Florbetapir ^18^F Positron Emission Tomography

^18^F-florbetapir (Amyvid, Eli Lilly) PET/CT data was collected from participants using a GE discovery MI digital scanner (Waukesha, WI, USA) according to the standardized procedures outlined by the Society of Nuclear Medicine and Molecular Imaging (3D acquisition; single intravenous slow-bolus < 10 mL; dose = 370 MBq; waiting period = 30–50 minutes; acquisition = 10 minutes) (45). The CT data was used to attenuation-correct the PET images, and the PET images were then reconstructed in MIMNeuro at a slice thickness of two millimetres, (46) converted to body weight-based voxel-wise standardized uptake values (SUVbw) and normalized into MNI space. A fellowship-trained neuroradiologist who was blinded to group assignment read and assessed each scan as being “Aβ positive” or “Aβ negative” based on established clinical criteria (46).

### Quantification of Mitochondrial Energetics from Peripheral Blood

Each participant provided a whole blood sample by venous puncture into EDTA tubes for quantification of mitochondrial respiratory function (Figure 1A). To isolate the mononuclear fraction, buffy coats from each sample were submitted to a Ficoll-Paque Plus (GE Healthcare) gradient centrifugation. Peripheral blood mononuclear cells (PBMCs) were then cryopreserved in Fetal Bovine Serum with 10% DMSO. Within six weeks of isolation the cells were thawed and underwent assessment. The Seahorse XF96 Analyzer (Seahorse Bioscience) was used to quantify oxygen consumption rate (OCR) of PBMCs via the mitochondrial stress test assay. PBMCs were plated at 500,000 cells per well and three OCR measurements were taken on 5-6 technical replicate wells. Measurements were taken prior to and upon injection of 3.5 μM oligomycin (Sigma; complex V inhibitor), 1 μM fluoro-carbonyl cyanide phenylhydrazone (FCCP; Sigma; mitochondrial oxidative phosphorylation uncoupler) and 14 μM rotenone + 14 μM antimycin A (Sigma; complex I and III inhibitors, respectively). This process aimed to evaluate features of mitochondrial respiration including basal respiration (pre-injection), proton leak (oligomycin), maximal respiration (FCCP), and non-mitochondrial respiration (rotenone and antimycin A). Each feature was calculated as the average of the three OCR measurements following each serial injection. Mitochondrial respiration associated with ATP production (ATP-linked respiration) was calculated as the difference between the average proton leak and the average basal respiration observed in the PBMCs. All bioenergetic data were treated as independent, continuous measures of mitochondrial function for subsequent analyses, and were normalized to protein in the well. Calculations were performed using Seahorse Wave software (v2.2.0).

**Figure 1.**
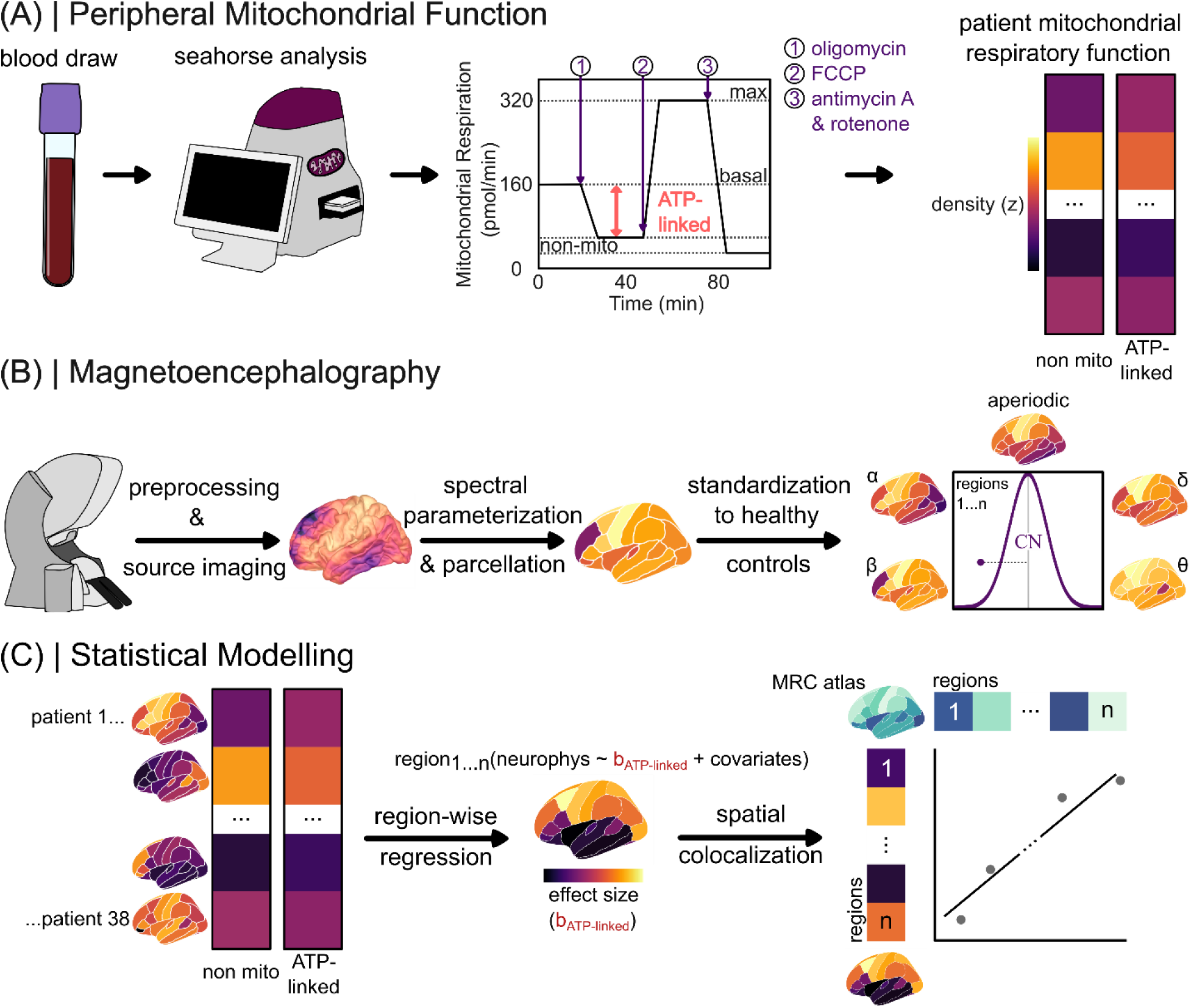
Study Design Elements. (**A**) A peripheral blood draw was taken from each participant, which was then placed in a Seahorse XF96 Analyzer. Mitochondrial respiration was taken continuously as various injections took place to replicate important measures of mitochondrial health including ATP-linked respiration. Individual participant mitochondrial respiratory function (non mitochondrial and ATP-linked) recorded. (**B**) Neurophysiological data was collected from each participant in the MEG, preprocessed and projected to the cortex. Spectral parameterization was then applied using *specparam* to separate rhythmic and arrhythmic signals, and the data was parcellated using the Desikan-Killiany atlas. These data were then standardized to z-scores using the maps from the control group. (**C**) Parcellated brain maps for each AD/aMCI participant were combined using region-wise regression to form beta-weight maps representing the overall correlation between ATP-linked respiration and altered neurophysiological signalling in each brain region, while accounting for covariates age, education, and non-mitochondrial respiration. The beta-weight maps were then compared to a map of mitochondrial respiratory capacity (MRC) (58) using spatial colocalization.

### Magnetoencephalography Data Collection and Processing

MEG data collection and processing closely follow that of previous publications (23,38,44,47–49; Figure 1B). To prepare participants for MEG measurement, four head position indicator coils were attached to the head and localized in addition to three fiducial points and about 100 scalp surface points using a Fastrak 3-D digitizer (Polhemus Navigator Sciences, Colchester, VT, USA). Participants were seated in a nonmagnetic chair and with their head positioned within the MEG sensor array, then asked to rest with their eyes closed for eight minutes (50). Recordings were conducted in a one-layer magnetically shielded room with active shielding engaged to compensate for environmental noise. Data was collected using a 306-sensor Elekta/MEGIN MEG system (Helsinki, Finland) equipped with 204 planar gradiometers and 102 magnetometers. Neuromagnetic responses were continuously sampled using an acquisition bandwidth of 0.1-330 Hz at a rate of 1 kHz. Each dataset was corrected for head motion and subjected to noise reduction using the signal space separation method with a temporal extension (correlation limit: .950; correlation window duration: 6 s) (51). All analyses were conducted using only data from the gradiometers. In addition to MEG data, T1-weighted structural MRI data (Siemens Prisma 3T; 64-channel head coil; repetition time [TR]: 2.3s; echo time [TE]: 2.98 ms; flip angle: 9°; field of view [FOV]: 256 mm; slice thickness: 1 mm; voxel size: 1 mm^3^) were collected for each participant to co-register their MEG data.

Triangulated cortical surfaces were derived from each participant’s T1 MRI data via *FreeSurfer* recon_all (52) using default settings followed by visual quality inspection. The triangulated surfaces were imported into *Brainstorm* (53), and each participant’s MEG data were co-registered with their MRI-derived data surfaces using digitized points of the scalp and fiducials via an iterative closest-point rigid-body registration in *Brainstorm*. This co-registration was manually corrected following a visual inspection. For MEG source imaging, individual cortical surfaces were down sampled to 15,000 vertices.

MEG data preprocessing and analysis was performed in accordance with recommended good-practice procedures (54). Once imported into *Brainstorm*, MEG data were filtered (bandpass = 1-200 Hz & notch = 60, 120, 180 Hz). An automated identification algorithm was used to identify ocular and cardiac artifacts followed by visual inspection of the artifact’s temporal and spatial topography. Signal-Space Projectors (SSPs) were generated for each type of artifact. The temporal and spatial topography of these SSPs were reviewed, and those accounting for ocular and/or cardiac components were removed from the data. MEG data were then epoched into four second non-overlapping blocks and down sampled to 500 Hz. Blocks that still contained major artifacts were excluded within each participant using the ∪ of standardized thresholds based on median absolute deviations (MAD) from the median (± 2.5 MAD) for peak-to-peak signal amplitude and gradient.

An overlapping-spheres forward model was used to source image MEG data. We used a linearly constrained minimum variance beamformer implemented in *Brainstorm* to spatially filter the epoch-wise data, with source orientations unconstrained to the cortical surface. Welch’s method (window = 1 s; 50% overlap) was used to transform the source-level time series data into the frequency-domain, and these spectra were then parameterized using *specparam* (55) (*Brainstorm* Matlab version; frequency range = 2 – 30 Hz; Gaussian peak model; peak width limits = 0.5 – 12 Hz; maximum *n* peaks = 3; minimum peak height = 3 dB; proximity threshold = 2 standard deviations of the largest peak; fixed aperiodic; no guess weight).

Six maps of neurophysiological brain activity were produced for each participant. The rhythmic features of the neurophysiological power spectrum were derived by subtracting the aperiodic components from the original PSDs and averaging over canonical frequency bands (delta: 2-4 Hz; theta: 5-7 Hz; alpha: 8-12 Hz; beta: 15-29 Hz). The arrhythmic features were derived by extracting the slope and offset of the aperiodic model fit at each vertex. Both rhythmic and arrhythmic features were then averaged within each region of the Desikan-Killiany atlas (56). This procedure produced four rhythmic maps, one for each canonical frequency band, and two arrhythmic maps (aperiodic exponent and offset). Here it should be noted that our analyses focused on the aperiodic exponent because the literature supports this measure specifically as a marker of excitatory-inhibitory balance (30,31). We then standardized these maps for each participant with aMCI and probable AD to the mean and standard deviation of the comparable maps from the cognitively normal control group, resulting in z-scored alterations in cortical neurophysiological signaling attributable to a biomarker-confirmed diagnosis of AD.

### Statistics

Linear models were used to test the relationship between ATP-linked respiration and neurophysiological changes in participants with Alzheimer’s disease using the lm toolbox in R (Figure 1C, left). ATP-linked respiration was modeled as a continuous variable with non-mitochondrial respiration, age, and education included as nuisance covariates. The final model took the following form:

Neurophysiological change ∼ ATP-linked respiration + non-mitochondrial respiration + age + education

For each neurophysiological feature of interest (delta: 2-4 Hz; theta: 5-7 Hz; alpha: 8-12 Hz; beta: 15-29 Hz, aperiodic exponent), this model was fit per region of the Desikan-Killiany atlas (56) and the resulting *p*-values associated with the effect of ATP-linked respiration were corrected across regions using the Benjamini-Hochberg method (57). Unthresholded maps of the effect size (i.e., beta weight) representing the association between ATP-linked respiration and neurophysiological changes were also extracted at this step for colocalization analysis (see *Alignments to a Cortical Atlas of Mitochondrial Respiratory Capacity*, below). All models were re-computed using an outlier exclusion threshold of normalized residuals ≥ the 97.5 percentile to confirm that our results were not influenced by outliers and only results that survived this post-hoc correction are reported.

### Alignments to a Cortical Atlas of Mitochondrial Respiratory Capacity

To test whether associations between cortical neurophysiological alterations and blood-based measurements of mitochondrial function were strongest in brain regions with high mitochondrial respiration, we contextualized these relationships using a human brain atlas of mitochondrial respiratory capacity created by Mosharov et al. (58). Briefly, this atlas is the result of a physical voxelization of a frozen human brain into 3 mm isotropic samples (58). Several measurements were taken from each of these voxels, including mitochondria-specific respiratory capacity and mitochondrial density, and the resulting maps were transformed into MNI space (58). We parcellated the resulting map using the Desikan-Killiany atlas (56) to facilitate comparison with the MEG data. We then conducted spatial colocalization analyses between the mitochondrial respiratory capacity atlas and brain maps of effect sizes from the region-wise linear models (described in the *Statistics* section) using the *cor.test* function in *R* and non-parametric spin tests with autocorrelation-preserving null models (5,000 Hungarian spins; threshold, *p*_SPIN_ < 0.05; Figure 1C, right) (59).

## Results

### Rhythmic and Arrhythmic Neurophysiological Alterations in Participants with Alzheimer’s Disease Pathology

We observed a general slowing of both rhythmic and arrhythmic neurophysiological activity in participants on the AD continuum, which was particularly pronounced in temporo-parietal cortices (Figure 2). Slow rhythmic activity in the delta and theta bands was increased in these participants relative to control levels, while alpha and beta frequency rhythms were decreased. There was also a steepening of the aperiodic slope (i.e., an increased exponent) in similar posterior cortical areas, indicating arrhythmic slowing.

**Figure 2.**
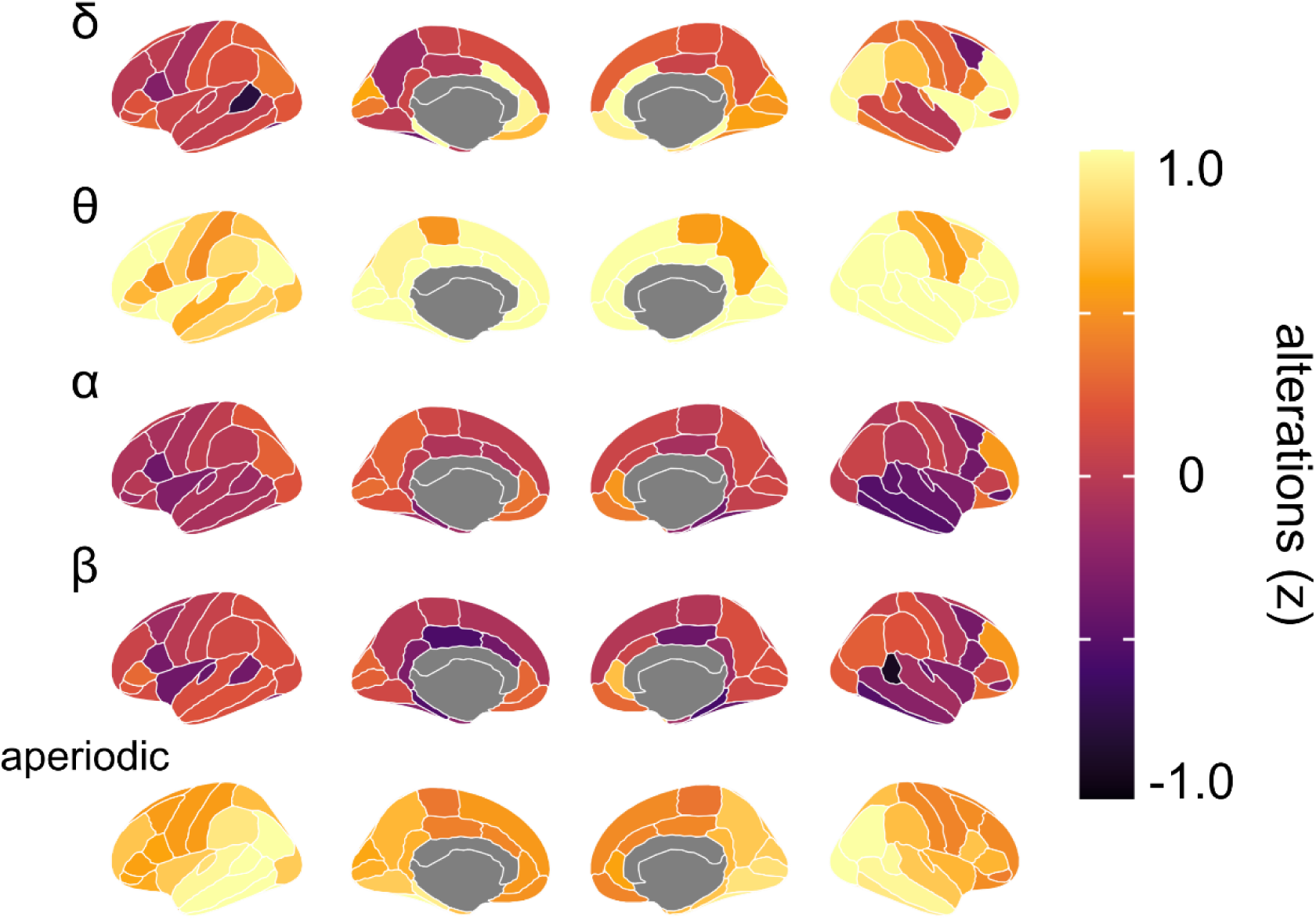
Rhythmic and arrhythmic neurophysiological alterations in participants with Alzheimer’s disease pathology. Brain maps indicate average alterations (in z-scored deviations from cognitively normal older adults) in activity across participants with Alzheimer’s disease pathology for each neurophysiological feature.

### ATP-linked Respiration is Associated with Altered Cortical Alpha and Theta Rhythms

We observed associations between blood-derived measures of ATP-linked respiratory capacity and rhythmic neurophysiological signaling (*p*_FDR_ < 0.05) in the theta (*r*_peak_ = -0.6; Figure 3A) and alpha (*r*_peak_ = -0.7; Figure 3B) bands. We found that reduced rhythmic signalling was associated with increased ATP-linked respiration across both bands (Figure 3A & B, right column). The theta associations were specific to the left middle temporal cortices (Figure 3A, left column), while the alpha effects were widespread, with the strongest associations in parieto-occipital cortices. These results survived outlier exclusion (removal of data points with normalized residuals ≥ the 97.5 percentile; *p*_FDR_ < 0.05). No significant associations were observed for other neurophysiological features.

**Figure 3.**
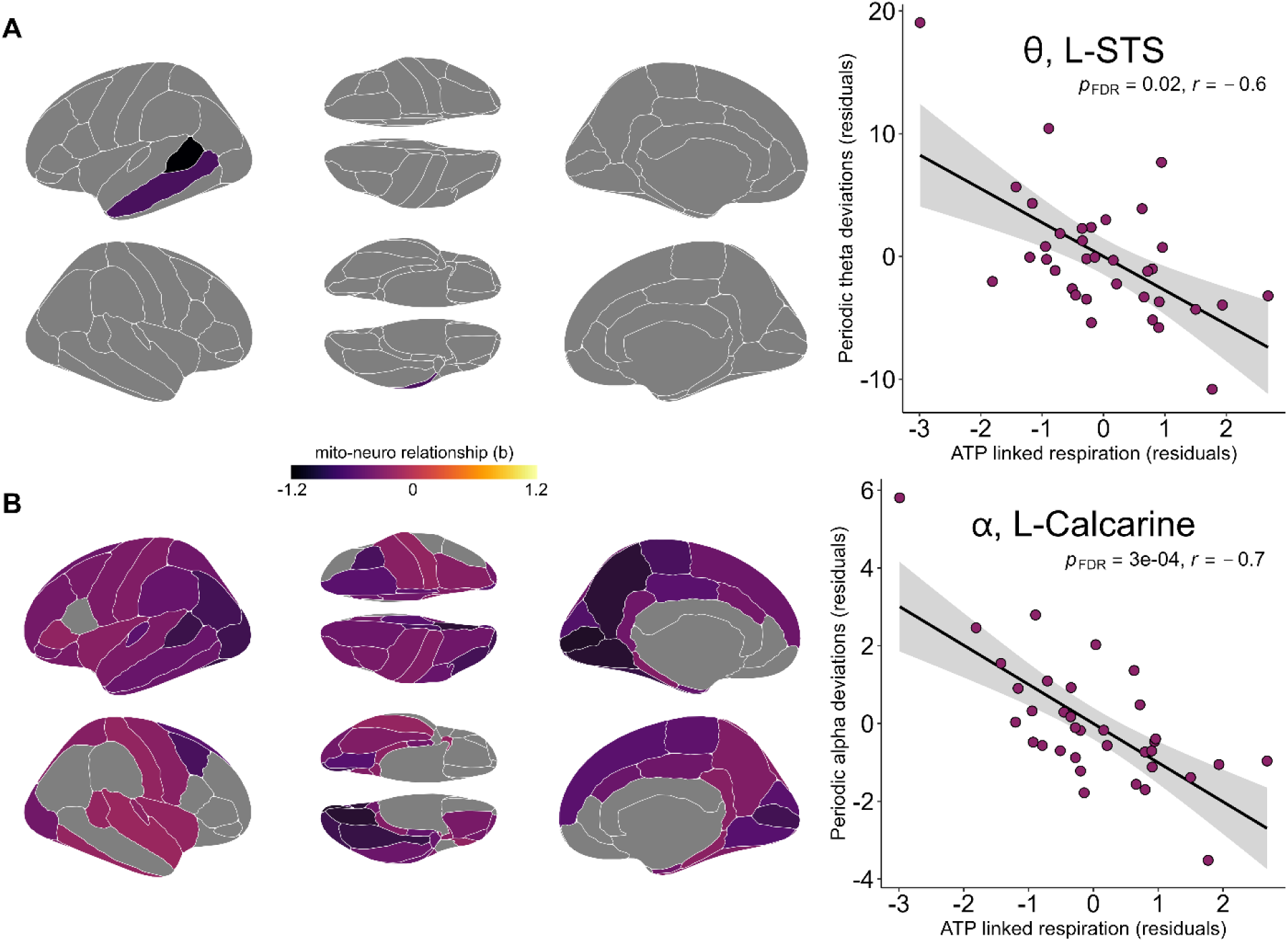
Associations between ATP-linked respiration and altered neurophysiological rhythms in participants with Alzheimer’s disease pathology. (**A**) *Left:* Brain-maps, parcellated using the Desikan-Killiany atlas, indicate regions in which the linear relationship between ATP-linked respiration and altered theta rhythms in AD was significant following correction for multiple comparisons (*p*_FDR_ < .05). *Right:* The scatterplot indicates the nature of this relationship at the region with the strongest effect, with periodic theta deviations from normal levels (y-axis; residuals; L-STS: left superior temporal sulcus) plotted against ATP-linked mitochondrial respiration (x-axis; residuals). (**B**) *Left:* Similar to (A), but for associations with periodic alpha-band deviations from normal levels. *Right:* Similar to (A), indicates the nature of the relationship at the region with the strongest effect (L-Calcarine: left calcarine sulcus).

### Subtle Alterations to Arrhythmic Neurophysiological Signaling in Brain Regions with Limited Mitochondrial Respiratory Capacity

Despite not reaching statistical significance in the region-wise analysis, we observed that associations between arrhythmic (i.e., aperiodic) neurophysiological activity and ATP-linked mitochondrial respiration exhibited a higher-order spatial alignment with a human brain map of mitochondrial respiratory capacity (58). Brain regions with reduced mitochondrial respiratory capacity were more likely to exhibit a relationship between lower ATP-linked mitochondrial respiration and aperiodic slowing (*p*_FDR_ = 0.003, *r* = 0.35; Figure 4). No significant relationships were found when we ran the same colocalizations using periodic components of cortical neurophysiological signalling.

**Figure 4.**
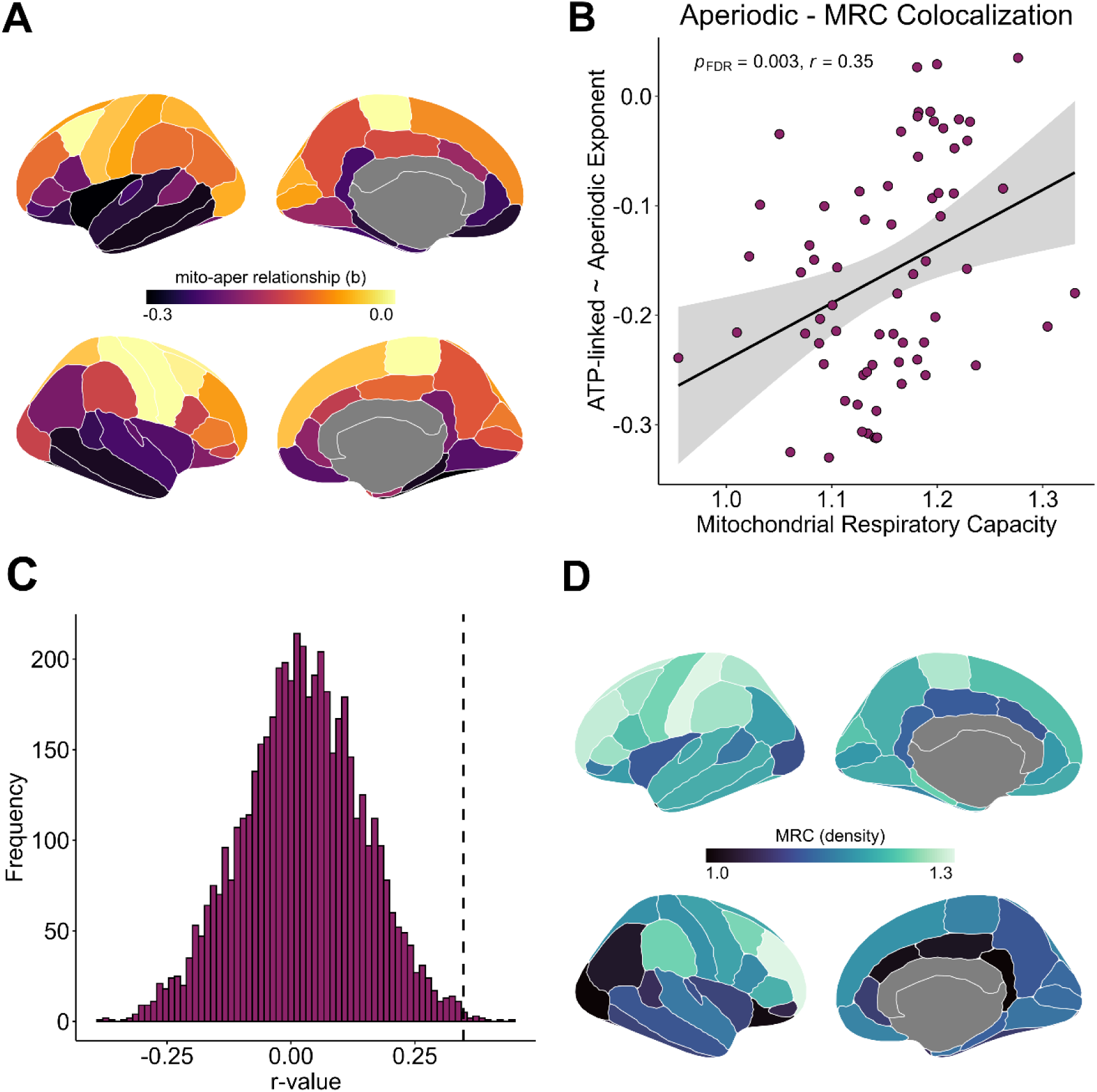
Topographical alignment of the relationship between ATP-linked respiration and alterations in aperiodic exponent with a human atlas of mitochondrial respiratory capacity. (**A**) Brain-maps, parcellated using the Desikan-Killiany atlas, indicate the regional strength of the correlation between alterations in aperiodic exponent and ATP-linked respiration. (**B**) The scatterplot indicates the colocalization between mitochondrial respiratory capacity (MRC) and the correlational strength between alterations in aperiodic exponent and ATP-linked respiration. Each dot in the scatterplot represents one Desikan-Killiany brain region. (**C**) The histogram plots the distribution of R-values observed during spin testing (x) against the frequency of each observation. The dotted line indicates our R-value’s position compared to those generated by chance during spin testing. (**D**) Brain-maps, similarly parcellated using the Desikan-Killiany atlas, indicate the mitochondrial respiratory capacity observed in each brain region. Lighter regions in this map indicate higher levels of mitochondrial respiratory capacity.

## Discussion

Alterations to both mitochondrial function and neurophysiological signalling have been implicated in AD. Despite strong evidence that these two variables are inextricably linked to one another in other clinical contexts, this relationship has not been examined in individuals on the AD continuum. We collected peripheral blood samples and task-free MEG from patients on the AD continuum, and used these data to elucidate the relationship between peripheral mitochondrial biomarkers and alterations in neurophysiological signalling in AD. We found that peripheral measures of mitochondrial function are correlated with rhythmic neurophysiological alterations across the theta and alpha frequency bands, as well as with subtler shifts in arrhythmic neurophysiology in cortical areas with lower respiratory capacity. These findings indicate that commonly reported alterations in brain signaling in patients with AD may be linked to mitochondrial dysfunction and suggest potential utility of peripheral mitochondrial assays for studying associations between energetics and neurophysiological changes in the context of age-related neurological disease.

Individuals on the AD continuum showed evidence of rhythmic neurophysiological slowing relative to controls, including reduced periodic power in the alpha and beta frequency bands, as well as increased power in the theta and delta bands. These patients also exhibited slowing of arrhythmic neurophysiology relative to controls. These alterations were most pronounced in temporo-parietal cortices and agree with previous literature on neurophysiological alterations in AD (3,22,30).

ATP-linked mitochondrial respiration, measured from peripheral blood draws, was associated with rhythmic neurophysiological alterations in both alpha and theta frequencies, such that increased respiration was associated with reduced signalling in both bands for the AD group compared to normal controls. Though supported by several human studies of AD (14–16), the mechanism and functional significance of increased mitochondrial respiration in these patients is unclear. We speculate that this may indicate a late-stage response to AD pathology, whereby the mitochondria begin to produce more ATP to compensate. In this context, and considering the established inverse relationship between metabolism and functionally-inhibitory cortical alpha oscillations (60–62), it is intuitive that a reduction of rhythmic cortical activity would accompany this change in mitochondrial respiration. The specificity of these associations to theta and alpha band activity is also intuitive, as rhythmic signalling in these frequencies is most consistently implicated in the hallmark cognitive declines (20,38,63–65) and proteinopathy (20,66) of AD.

Despite evidence suggesting that peripheral assays can effectively predict mitochondrial respiratory function in the brain (17,21), the absence of direct measures of brain mitochondria limits our interpretations. To partially address this limitation, we compared the spatial topographies of associations between signaling changes and peripheral mitochondrial function to a cortical map of mitochondrial respiratory capacity (58). Surprisingly, we found that none of the strong associations between peripheral ATP-linked respiration and rhythmic cortical signaling were aligned to this atlas of mitochondrial respiratory capacity. This is potentially due to the generation of theta-alpha oscillations via thalamo-cortical loops originating in thalamic reticular nuclei (67,68). In other words, alterations in theta and alpha cortical rhythms in AD are more likely to be attributable to mitochondrial dysfunction in thalamic nuclei than in the neocortex. Supporting this concept, we found that more subtle slowing of arrhythmic neurophysiology – thought to index local excitatory-inhibitory balance (69–71) – was associated with mitochondrial dysfunction more strongly in regions of the brain with lower mitochondrial respiratory capacity.

Our study has several limitations that should be considered. First, the absence of direct measures of mitochondrial function makes it difficult to directly link peripheral measures of mitochondrial function to macro-scale neurophysiology while excluding all potential mediating factors. This limitation has been addressed to some degree by previous research indicating peripheral mitochondrial assays as a less invasive predictor of bioenergetics in the human brain (17, 21). Second, it is also important to note that the Mosharov atlas used for spatial contextualization of our results only measured mitochondrial function in the brain of one individual. This limits the generalizability of these findings somewhat, but represents the best resource currently available for such an analysis. Future studies generating similar atlases from multiple individuals, or benchmarking the existing atlas against sparser samples of brain tissue from a larger group, would help advance the field. Another limitation of our study includes the lack of data from individuals in the pre-clinical stages of AD. The pathophysiological process of AD begins many years before onset of clinical symptoms, and future research linking peripheral mitochondrial function to neurophysiological signalling in asymptomatic adults with AD proteinopathy would be incredibly valuable. Finally, while the sample size of our study is larger than those typical in this field (72), it is not large enough to allow for assessment of sex effects, clinical subtypes, atypical disease variants, and key clinical features that may moderate the reported findings. It would be valuable to replicate this study with a larger population to fully capture the heterogeneity of AD.

Our findings indicate a link between mitochondrial dysfunction in AD and rhythmic alpha-theta signalling alterations, potentially originating in thalamic nuclei, as well as with arrhythmic activity originating in the cortex. This study provides further evidence of the proposed link between mitochondrial dysfunction and neurophysiological signalling, and extends this literature into patients with AD.

## Data Availability

Due to concerns over patient confidentiality the MEG and PET data from the DMAP study cannot be shared on an open repository, but will be made available from the corresponding author upon reasonable request.

## Acknowledgements

This work was supported by the Canada Research Chair (CRC-2023-00300) in Neurophysiology of Aging and Neurodegeneration and a Discovery Grant from the Natural Sciences and Engineering Research Council of Canada (RGPIN-2025-04783) to AIW, as well as grants from the National Institutes of Health of the United States (R01-MH116782 and R01-MH118013) to TWW.

